# Assessing the Compliance and Timeliness of Results Reporting for Clinical Trials on Antimicrobial Agents

**DOI:** 10.64898/2025.12.23.25342922

**Authors:** Megan Curtin, Allisun Wiltshire, Gustav Nilsonne, Maximilian Siebert

## Abstract

**Objective:** Antimicrobial resistance (AMR) is an urgent global health threat, resulting in more than 5 million deaths globally in 2019. Timely and complete antimicrobial agent (AMA) clinical trial results reporting is essential to evaluate the safety and efficacy of investigational therapies. The Food and Drug Administration Amendments Act (FDAAA) of 2007 mandated results reporting for applicable clinical trials to ClinicalTrials.gov. After nearly ten years of underreporting, the HHS issued the Final Rule, requiring a designated responsible party to submit results to ClinicalTrials.gov and clarifying applicable clinical trial (ACT) criteria. ACTs and probable ACTs (pACTs) are interventional studies regulated by the FDA with at least one site based in the United States. However, pACTs were initiated prior to January 2017, when the Final Rule came into effect. This study investigates the compliance and timeliness of results reporting of ACTs and probable ACTs (pACTs) for AMAs.

**Design:** We extracted data from ClinicalTrials.gov for trials involving AMAs with primary completion dates between May 1, 2013, and May 1, 2023. We analyzed the time from primary completion to results reporting and estimated the hazard ratio to compare timeliness between ACTs and pACTs. Additionally, we assessed delays in reporting across different study types and funding sources.

**Results:** Our search resulted in 2629 NCT records. After exclusion of ineligible trials, we included 2525 trials. We found 1769 pACTs (70.1%; 95% CI, 69.3%-72.9%) and 756 ACTs (29.9%; 95% CI, 28.2%-31.8%). Among the 2525 eligible trials, 2249 trials (89.1%; 95% CI, 87.8%-90.2%) were reported on ClinicalTrials.gov or in journal publications. Overall, 81.3% (95% CI, 79.7%-82.3%) of trials were reported late or missing (75.0% of ACTs vs 83.6% of pACTs). ACTs were more likely to report results earlier than pACTs, with a hazard ratio of 1.4 (95% CI, 1.3-1.5).

**Conclusions:** ACTs demonstrated greater reporting compliance and shorter delays in the reporting of overdue results. While this analysis provides initial insights, limitations related to timeline and sample scope suggest that broader investigations are needed to fully evaluate the impact of the Final Rule.

## Introduction

Antimicrobial resistance (AMR) has emerged as a critical global health threat and is now recognized as one of the leading causes of death worldwide, with the greatest burden falling on low- and middle-income countries (LMICs). ^1^ The United States is not exempt from this trend. In 2013, the U.S. National Action Plan for Combating Antibiotic-Resistant Bacteria reported an estimated 2 million people were infected with antibiotic-resistant bacteria and 23,000 individuals die annually from these pathogens.^2^ The U.S. Center for Disease Control (CDC) found in 2019 that these figures increased, with an estimated 2.8 million antibiotic-resistant infections and more than 35,000 deaths annually. Beyond the human toll, AMR also imposes a significant economic burden: American taxpayers pay $4.8 billion in medical expenses to treat only a fraction of these AMR infections.

Research and innovation in the AMR space are essential to generate efficacious medicines. AMR occurs in a variety of microorganisms such as bacteria, viruses, fungi, and parasites that have evolved mechanisms to evade susceptibility to existing antibiotics, antivirals, antifungals, and antiparasitics. In 2021, only six antimicrobial agents in clinical development met at least one of the World Health Organization’s criteria for innovation. Barriers to the development of novel therapies are exacerbated by emerging resistance occurring on average 2-3 years post-market entry for new antimicrobial products.^3^

Timely results reporting for AMR trials is essential to ensure these medicines are safe and effective for public usage. Analysis of antimicrobial agent research quality and results reporting could also ameliorate limitations in therapeutic innovation. However, there is minimal literature on the timeliness and compliance of clinical trial results reporting for antimicrobial agents. Historical underreporting of trial results has led to notable cases of patient harm. The United States spent $1.7 billion stockpiling Tamiflu (Oseltamivir), an antiviral for the H1N1 pandemic.^4^ Despite many RCTs investing Tamiflu, undisclosed serious adverse events resulted in cardiac arrhythmia and suicidal ideation in children.^5^ The example of Tamiflu demonstrates the need for timely and complete results reporting to ClinicalTrials.gov to not only ensure a fair return of taxpayer investment, but to also safeguard public health. Understanding how these obligations apply to antimicrobial trials requires examining the FDA’s framework for results reporting, particularly the classifications of Applicable Clinical Trials (ACTs) and probable ACTs (pACTs).^6^

Applicable Clinical Trials (ACTs) are interventional studies other than phase 1 and are regulated by the U.S. FDA with at least one site based in the U.S. ACTs have a primary completion date on or after January 2008 when FDAAA was initiated and a study start date after the Final Rule implementation in January 2017. Probable Applicable Clinical Trials (pACTs) adhere to the same criteria but have a study start date before the “Final Rule” for Clinical Trials Registration and Results Information Submission came into effect. ^7^ The Final Rule requires ACTs to designate one responsible party for ClinicalTrials.gov results reporting, while also mandating the Secretary of Health and Human Services to use rulemaking to improve results reporting on ClinicalTrials.gov under FDAAA Section 801. As of November 2025, the United States government could have imposed fines of up to $100 billion due to underreporting of clinical trial data, with 21.8% of all clinical trials registered on ClinicalTrials.gov with outstanding results.^8^

Given the current gaps in compliance, it is essential to understand how clinical trials involving AMAs perform under these regulatory obligations. Hence, this study aims to investigate the compliance and timeliness of results reporting of ACTs and pACTs for antimicrobials.^7^ The ACT/pACT distinction is particularly important for AMA trials, as pACTs reflect legacy reporting practices while ACTs fall under the fully implemented Final Rule, an issue of consequence given how rapidly resistance trends change and how essential timely evidence is in AMR research. Moreover, we analyze how the characteristics and reporting for antimicrobial trials varies based upon ACT or pACT designation.

## Methods

We utilized ClinicalTrials.gov to identify AMA trials completed over a 10-year period between May 1st, 2013 and May 1st, 2023. This timespan ensures more than one year of follow-up for assessing compliance with the federal reporting deadline. Every study registered in this database receives a unique National Clinical Trial (NCT) identifier, therefore we were able to track each trial consistently across registry fields and publication sources. We measured the time from the primary completion date to results submission and calculated hazard ratios to evaluate differences in reporting timeliness between ACTs and pACTs. As defined in the ClinicalTrials.gov database, the primary completion date is the date on which the last participant in the study was examined or received an intervention to collect final data for the primary outcome measure. The federal results reporting deadline is one year after the primary completion date.^7^ We also captured the time delay in results reporting across study designations, funding sources, phases, and product type.

Our search strategy for ClinicalTrials.gov can be found in Box 1.

### Box 1. Search Strategy

**Check ClinicalTrials.gov for:**

a. Intervention: ‘Anti-infective’ OR ‘antimicrobial’ OR ‘antibiotic’OR ‘antibacterial’ OR ‘antiviral’ OR ‘antifungal’
b. Study Type: Interventional
c. Location: United States
d. Primary Completion Date: May 01, 2013 to May 01, 2023
e. Phase 2/3/4
f. Study Results: With Results, Without Results
g. Study Status:
  i. Looking for participants (exclude not yet recruiting)
    1. Recruiting
  ii. No longer looking for participants
    1. Active not recruiting
    2. Completed
    3. Terminated
  iii. Other (exclude withdrawn)
    1. Enrolling by invitation
    2. Suspended
    3. Unknown
h. Download the aggregate analysis of ClinicalTrials.gov (AACT) data set including the following fields:
  i. NCT number, Study Title, Study URL, Study Status, Conditions, Interventions, Primary Outcome Measures, Sponsor, Collaborators, Phases, Enrollment, Funder Type, Study Type, Study Design, Start Date, Primary Completion Date, Completion Date, First Posted, Results First Posted, Locations

Eligibility requirements included trials that were not phase 1, not testing for medical device feasibility, only interventional, and with at least one site in the United States.

### Manual Data Extraction for Publications

We analyzed the cohort of NCTs with outstanding results, as of the extraction date, November 15 2024, to evaluate whether results were reported in publications. Two independent researchers (MC & AW) searched the ClinicalTrials.gov publication section. If no publications were listed, we entered the NCT number into Google and screened the first page. If this process did not yield a publication, we performed another Google search with two combinations of terms from the registry (e.g., title, principal investigator, intervention, disease, or other). We matched each completed trial to articles indexed in PubMed using a structured set of criteria to identify corresponding peer-reviewed publications. We first verified the ClinicalTrials.gov identifier (i.e. NCT number) reported in the publication whenever available. If no NCT number appeared with this search criteria, we matched publications based on concordance of key trial descriptors, including the intervention, indication, study design, and sample size. We required that the publication reported primary clinical results from the trial, and excluded articles that were not original research (e.g., reviews, commentaries, methodological papers). To ensure temporal validity, the publication date had to follow the start date of the trial. When multiple candidate publications existed, the earliest publication was retained. Publication dates were extracted automatically through PubMed (for articles with PubMed IDs) and otherwise manually from the publication, preferring the earliest date among multiple available dates or eligible publications.

### Statistical Analysis

All statistical analyses were conducted in R. Cumulative percentages and time-to-event outcomes were assessed using the Kaplan–Meier method. When the date of results publication preceded the primary completion date, we input the publication date as the primary completion date for the trial.

To examine the association between trial type (ACTs and pACT) and results availability over time, hazard ratios (HRs) were estimated using Cox proportional hazards models. All P values were two-sided, and estimates were reported with 95% confidence intervals (CIs). A significance threshold of P < .05 was applied to all analyses.

Our pre-registered protocol along with deviations can be found here: https://osf.io/zp8ug/overview.

## Results

Our initial search generated 2793 trials. After screening trials that failed to meet ACT and pACT criteria, we excluded 164 trials. We examined the remaining 2629 trials to further remove 104 NCTs that only included agents with nonantimicrobial properties. As a result, we identified 2525 eligible trials (Figure 1). A median enrollment of 60 patients (IQR, 25-207) was found. Within this sample, there were 1769 trials identified as pACTs (70.1%; 95%CI, 69.3%-72.9%) and 756 as ACTs (29.9%; 95%CI, 28.2%-31.8%). NCTs with outstanding results demonstrate a delay of nearly four years (3.8 years) from the federal deadline.

**Figure 1:**
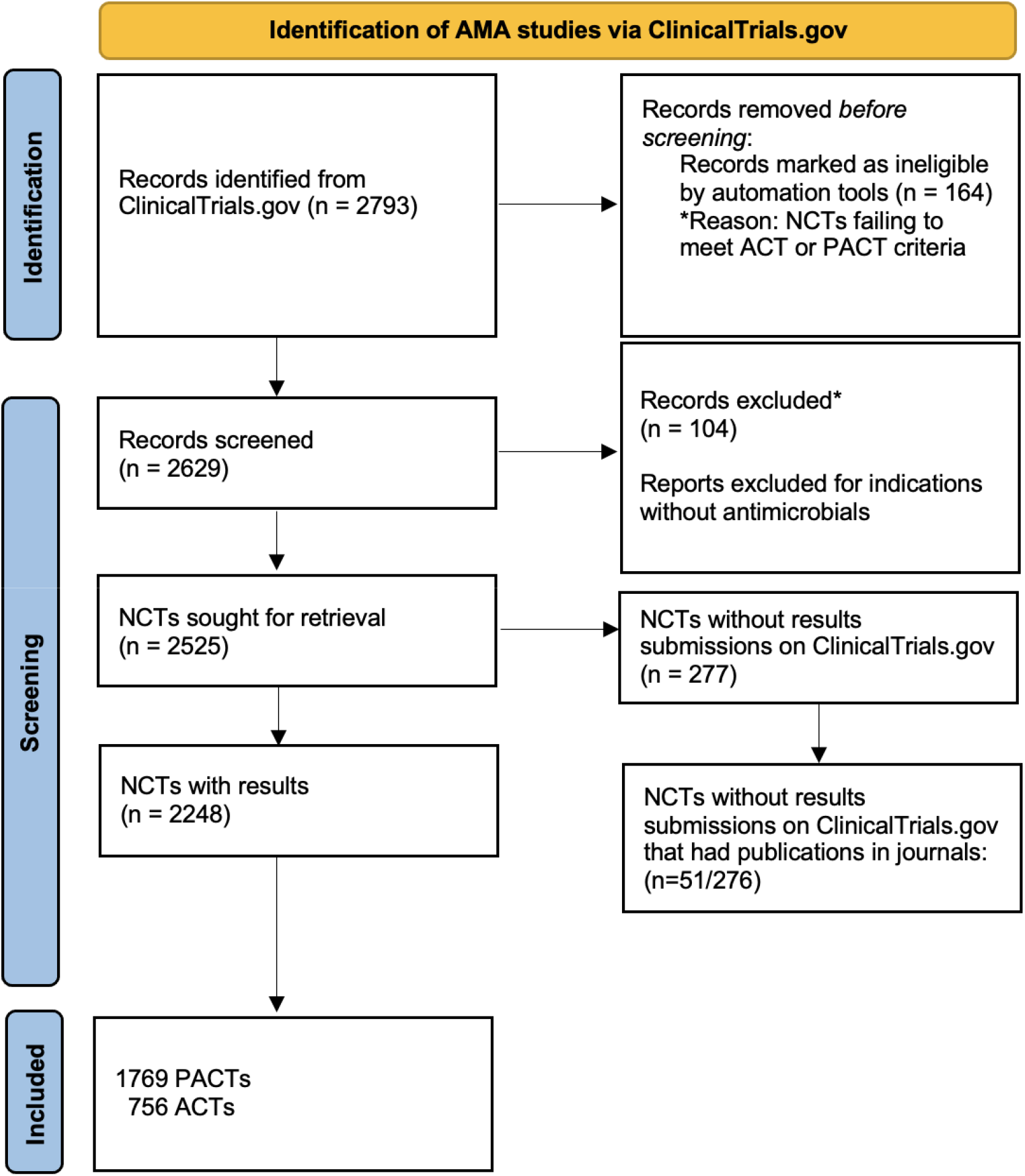
Identification of AMA Studies via ClinicalTrials.gov.

Regarding the trial characteristics, Phase 2 trials represent the largest proportion (45.3%), followed by Phase 3 (24.0%) trials (Table 1). The antimicrobial agent interventions are primarily involved in cancer-related trials (36.4%) and addressed viral diseases (25.4%). The cancer-related trials predominantly evaluated antimicrobial agents prescribed for opportunistic infections in immunocompromised oncology populations. Regarding therapeutic focus, 36.4% of the interventions were used in trials with cancer indications and 25.4% targeted viral diseases. The primary outcome measure in these trials not only included infection treatment, but also tumor response. Funding sources were diverse, most of the trials, 37.8%, were industry-funded, followed by 22.9% that were sponsored by universities and just over half of all studies (57.8%) were randomized.

**Table 1:** Characteristics of Clinical Trials in Sample.

| Characteristics | All Trials<br>N=2525, (%) |
| --- | --- |
| ACT | 756 (29.9) |
| pACT | 1769 (70.1) |
| **Trial Phases<sup>1</sup>** |  |
| Phase 1/2 | 298 (11.8) |
| Phase 2 | 1143 (45.3) |
| Phase 2/3 | 80 (3.1) |
| Phase 3 | 605 (24.0) |
| Phase 4 | 399 (15.8) |
| **Antimicrobial Agent Type Use** |  |
| Bacterial | 233 (9.2) |
| Cancer | 919 (36.4) |
| Generalized infection | 69 (2.7) |
| Viral | 639 (25.3) |
| Other<sup>2</sup> | 665 (26.4) |
| **Funding Types** |  |
| University | 577 (22.9) |
| Government | 314 (12.4) |
| Industry | 953 (37.7) |
| Hospital System | 335 (13.3) |
| Individual | 54 (2.1) |
| Other | 293 (11.6) |
| **Randomized** |  |
| Yes | 1459 (57.8) |
| No | 1066 (42.2) |

<sup>1</sup> Phase 1 was excluded due to non-obligation to register Clinical Trials
<sup>2</sup> Ophthalmologic, dermatologic, behavioral, renal disease, transplant rejection, cardiovascular disease, diabetes, genetic disorder, inflammatory disorder

In terms of results reporting, 2249 of the 2525 sampled trials (89.1%; 95% CI, 87.8%–90.2%) had results posted on ClinicalTrials.gov, of which 70.1% were pACTs and 29.9% were ACTs. As of November 15, 2024, 2053 of these 2249 trials (91%) reported results after the one-year federal deadline. Among the 276 trials without posted results, we identified 51 publications, bringing the total number of trials with results available either on ClinicalTrials.gov or in a publication to 2301 (91.1%).

With regards to timeliness, NCTs with publications had a median delay (IQR) of 0.3 years (0.2-0.8). There was a median (IQR) of 0.5 years (0.2-1.5) between 1 year after the primary completion date and the results submissions across all studies. The median (IQR) time lag for late reporting for ACTs was 0.3 years (0.1-0.9), while it was 0.6 years (0.2-1.8) for pACTs. Overall, 81.3% (95% CI, 79.7%-82.3%) of trials were reported late or missing (75.0% of ACTs vs 83.6% of pACTs).

Figure 2 displays the cumulative incidence of results reporting among ACTs and pACTs from the primary completion date through 12 years of follow-up. At 1 year after the primary completion date, 26.7% of ACTs had reported results compared with 17.1% of pACTs, indicating substantially faster early reporting among ACT-designated trials. After 6 years, ACTs showed 96.7% results reporting while 90.5% of pACTs had submitted results. The difference between the two groups was statistically significant (log-rank P < .001). In the Cox proportional hazards model, ACT designation was associated with a 40% higher hazard of results reporting compared with pACTs (HR, 1.4 [95%CI 1.3-1.5]), indicating that ACTs were reported more frequently and earlier.

**Figure 2:**
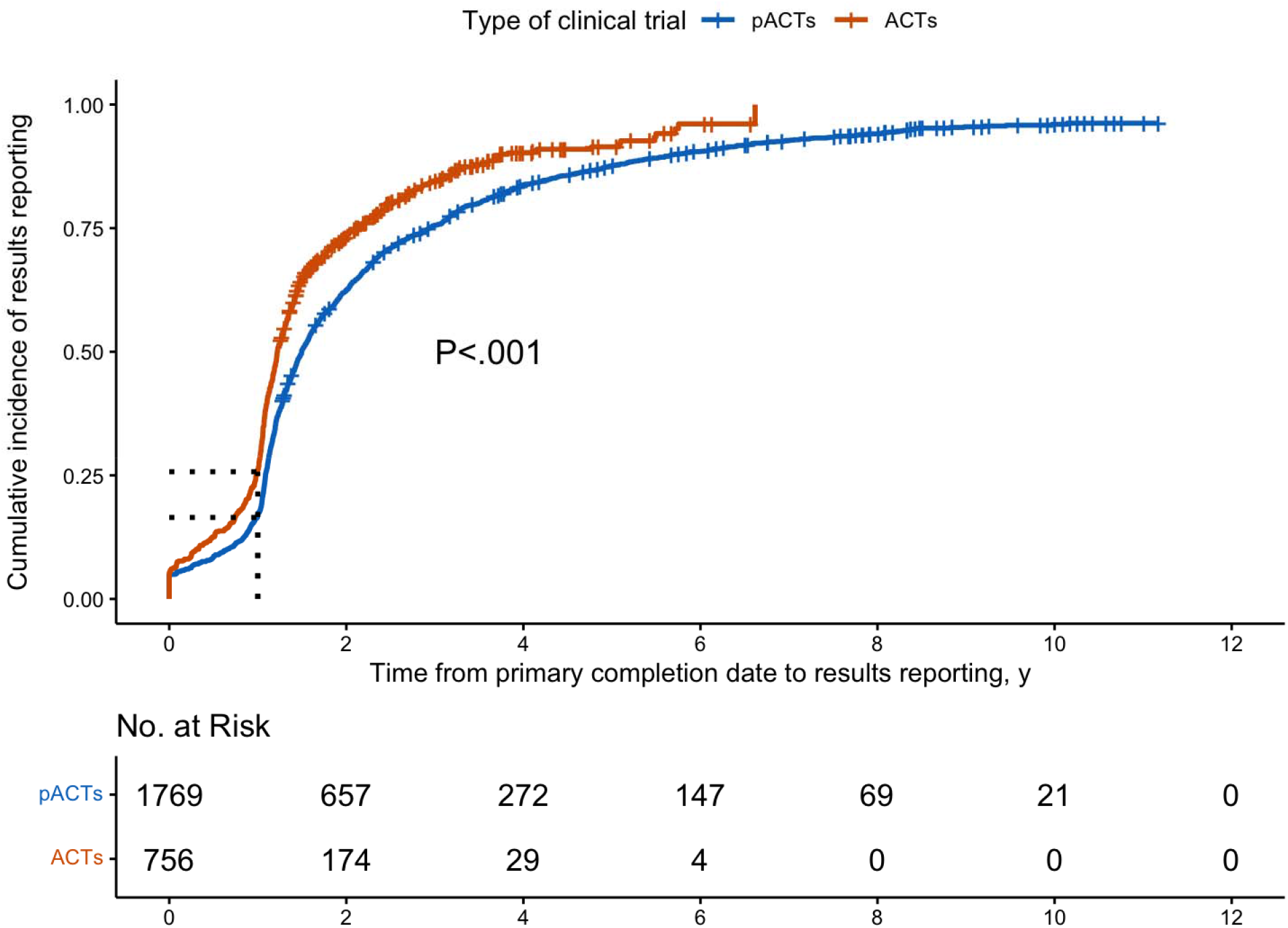
Kaplan-Meier Plot for all, published on ClinicalTrials.gov or in Publication. The curves show the cumulative incidence of reporting from the primary completion date through 12 years of follow-up, stratified by Applicable Clinical Trials (ACTs) in orange and probable ACTs (pACTs) in blue. The vertical dashed line at 1 year represents the federal reporting requirement under FDAAA 801, which mandates that results be submitted within 12 months of the primary completion date A log-rank test demonstrated a statistically significant difference in time-to-reporting distributions between ACTs and pACTs (p < .001).

Regarding investigational areas, trials with viral indications reported the most results(92.4%), whereas NCTs involving generalized infections demonstrated the least submissions with only 81.1% of trials reporting results. Trials with cancer indications had the slowest reporting (median [IQR] delay, 0.6 [0.2-1.2] years), whereas viral and generalized infection trials had the most timely reporting (0.4 [0.2-1.0] years and 0.4 [0.2-1.2] years, respectively). (Table 2)

**Table 2:**
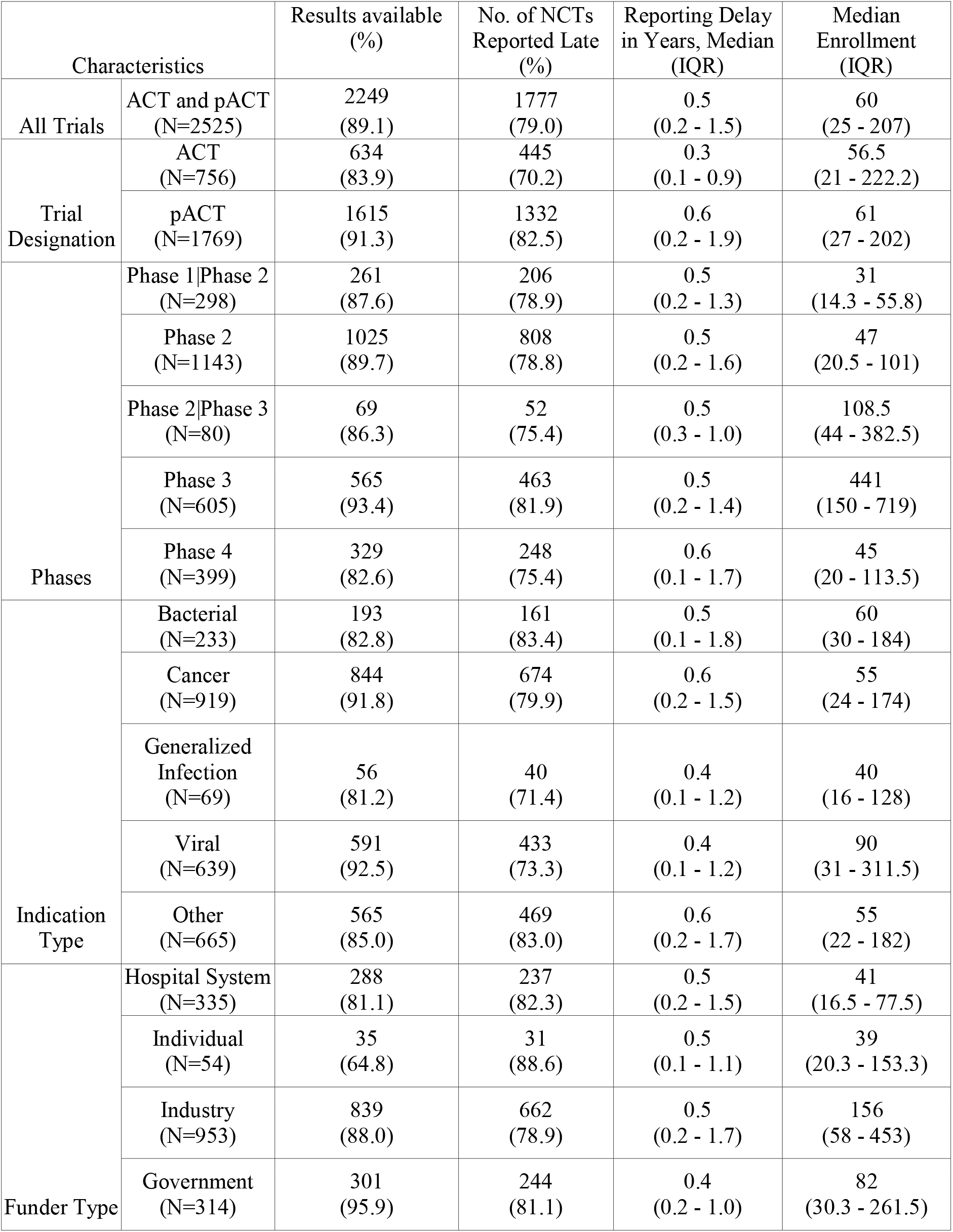
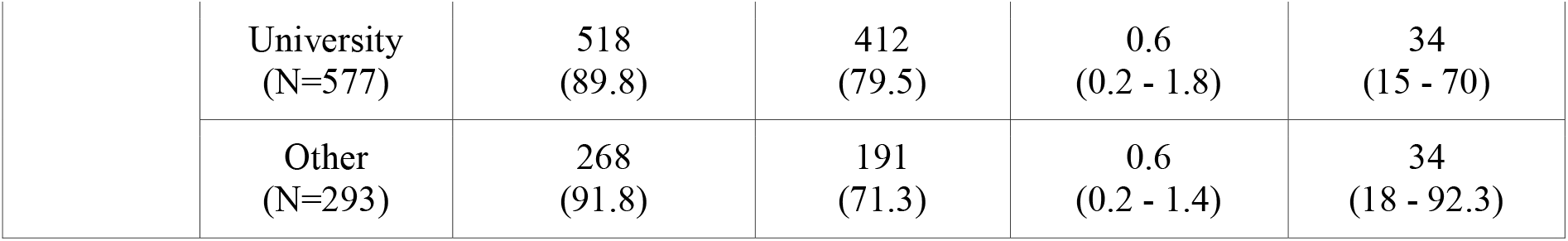
Late Results Reporting by Trial Characteristics on Clinicaltrials.gov.

Regarding funding sources, government-funded trials reported the most results with 96.4%, whereas trials with funding from individuals such as foundations reported the least trials with 64.8%. Universities had the slowest reporting (0.6 [0.2-1.4] years), while government-funded trials had the most timely reporting (0.4 [0.2-1.0] years).

## Discussion

### Main results

In this cross-sectional analysis of 2525 antimicrobial clinical trials registered on ClinicalTrials.gov, we found substantial delays in results reporting. Only 21% of trials reported results by the federal deadline and the median delay among late-reporting trials was nearly four years. ACTs reported results significantly faster than pACTs, with a hazard ratio of 1.4, consistent with influence of post–Final Rule regulatory obligations on reporting behavior. Reporting timeliness also varied by funder type: private donor–funded trials reported results with the greatest delays, whereas government-funded trials demonstrated the shortest delays. Among the antimicrobial agent NCTs with late results reporting, viral agents were the ones with the highest reporting rate and also had the shortest median delay of 0.3 years. AMAs used in oncology trials showed the highest percentage in late reporting. Furthermore, many results remain missing revealing persistent gaps in transparency for antimicrobial research and raising concerns about the timely availability of safety and efficacy data.

### Comparison with earlier research

Despite showing higher rates of overall reporting, the findings in our sample rates fall behind the standard reporting timelines on ClinicalTrials.gov. Whereas DeVito et al found in an overall sample of Clinicaltrials.gov 63.8% percent of trials reported, they identified 40.9% of randomized NCTs having reported results by the federal deadline.^9^

When comparing specific areas, Pellat et. al investigated clinical trial results reporting in NCTs for pancreatic cancers. Similar to our analysis, Pellat et al. found more timely results reporting for ACTs. However, 70% of results were posted either through publication or registration on ClinicalTrials.gov, while our team found 91% (2304/2525) of AMR NCT results reporting across publications and registration on ClinicalTrials.gov. ^10^ The reporting difference may be due to Pellat et. al having investigated results submissions for NCTs related to pancreatic adenocarcinoma, a specific kind of cancer, rather than across a spectrum of oncology trials. Trials with cancer-related indications in our study also demonstrated the slowest reporting with a median delay of 0.6 years, while viral AMAs had the fastest reporting time with a median delay of 0.4 years. Although this is a small difference in reporting, it may be a result of viral trials involving fewer data and shorter timeframes that yield improved reporting compliance by the federal deadline. Oncology trials typically last longer than other trial types.^11^ As a result, extensive NCT data analysis in trials with cancer indications may impact the feasibility of results reporting by the federal deadline on ClinicalTrials.gov. This observation aligns with prior research showing that oncology trials are among the slowest to report results. In a study of 10,422 cancer trials, only around 18% had posted results on Clincialtrials.gov and a small proportion, 6%, reported results in a publication.^12^

In our sample, universities had the greatest delays in AMR results reporting reflecting similar findings in other literature that academic centers demonstrate the poorest compliance.^13^ These institutions typically have fewer resources than industry sponsors which may contribute to deficits in reporting rates. For example, Emam et al. found that trials funded by academic institutions were less likely to use an electronic data capture system compared to industry-funded trials.^14^ These systems expedite data submissions and monitoring which may explain the deficits in academic center reporting rates. Moreover, government-funded studies had the fastest reporting timelines, which is contrary to other literature, with industry sponsor typically having the fastest reporting rates. Industry-sponsored trials also had lower compliance rates than university and government-funded trials.^13^ One possible explanation is that AMR-focused companies operate in a small and commercially fragile market, creating incentives to delay public disclosure to preserve competitive advantage or extend perceived exclusivity around investigational products.

ACTs demonstrate more timely results reporting compared to pACTs, suggesting the Final Rule generates increased compliance with FDAAA, and a general trend over time towards improved reporting. However, the proportion of Clinical Trials with compliant results has not improved since July 2018.^9^ Devito et al. maintain that greater enforcement of FDAAA is needed to elicit increased clinical trial results reporting. Nonetheless, recent analyses underscore the persistent challenges in clinical-trial transparency despite regulatory reforms. Mughal et al. found that following implementation of the Food and Drug Administration Amendments Act (FDAAA) Final Rule in 2017, 12-month reporting rates on ClinicalTrials.gov increased from 8.3% (2010– 2014) to 23.2% (2017–2021), and 36-month rates rose from 40.8% to 49.2%, yet the vast majority of trials remained out of compliance.^15^ A subsequent sponsor-level study revealed considerable variation: among 10,605 applicable trials completed 2017–2024, overall 12-month compliance averaged just 37.2%, with industry sponsors achieving 73.7%, NIH 71.0%, and academic sponsors only 25.5%.^13^

In a cross-sectional analysis of the European Union Clinical Trials Registry and ClinicalTrials.gov, Nilsonne et al. found the median time from completion date to results reporting to be 1.8 years whereas we found a median delay of 0.5 years for AMR trial results reporting. The reporting delays identified in the EUCTR may be related to differences in regulations. FDAAA requires results reporting within 12 months, while the team led by Nilsonne analyzed trials completed before the European Medicines Agency initiated the requirement for results reporting within 12 months for trials launched after January 2022 to the Clinical Trial Information System, a central reporting database for all EU trials.^16–18^

Together, these findings suggest that while regulatory changes may have produced modest improvements in results-reporting rates, systemic non-compliance persists and is strongly influenced by sponsor type, which has meaningful implications for trial transparency, research integrity, and public trust.

### Limitations

This study has several limitations. Although FDAAA applies only to ClinicalTrials.gov and does not govern reporting in other registries, our objective was to evaluate the reporting patterns of antimicrobial trials specifically within this U.S. registry; as a result, results disclosed exclusively in other national or international registries or published without being linked in ClinicalTrials.gov may not have been captured. Furthermore, ACT and pACT status is inferred using publicly available fields and may be misclassified in some cases. Publication searches may have also missed articles not indexed in Google or PubMed, and we did not assess the completeness or accuracy of the results that were reported. Similarly, the ClinicalTrials.gov search criteria could be tailored to automate the exclusion of nonantimicrobial agents, to reduce manual verification. The search terms were specified for AMA intervention types, yet could also include the indication types to broaden the results. Finally, we did not evaluate the quality of the posted results, which is an important dimension of trial transparency.

### Implications and Policy Recommendations

Our results show that despite high results submissions in specific AMA intervention areas, gaps remain. Underreporting of trial results contributes to redundancy and inefficiency in research. Without access to prior trial data, researchers may unknowingly replicate studies, exposing participants to unnecessary risk and wasting scarce resources. This problem is particularly acute in AMR research, where patient populations are often small and clinical trial opportunities are limited. The lack of publicly available results impairs clinical decision-making as well. Physicians and public health authorities depend on complete and timely data to guide evidence-based treatment decisions. Delays or omissions in trial reporting can result in suboptimal prescribing, inappropriate use of antimicrobials, and accelerated development of resistance, ultimately affecting patient outcomes. Safety monitoring is another area impacted by limited reporting. Adverse events or unexpected safety signals may remain undisclosed, putting current and future patients at risk. This concern is particularly pressing for novel AMR agents, which often have limited post-marketing experience and rely heavily on published trial data to establish safety profiles.

The importance of timely and complete reporting is underscored by a recent review showing that nearly 40% of antibiotic trials did not report any adverse events. When fundamental safety data are missing, clinicians and regulators cannot accurately evaluate risk, leading to potential patient harm and misguided treatment decisions. This illustrates that delays or omissions in AMR trial reporting directly weaken the evidence base needed for safe antimicrobial use.^19^Although our study focuses on reporting of primary outcomes based on primary completion dates, these timelines are closely linked to the availability of safety information, which is essential for responsible clinical and regulatory decision-making.

To counter this, the FDA and NIH have developed a range of training seminars and educational resources to improve compliance with ClinicalTrials.gov reporting requirements and enhance user proficiency. Yet, several opportunities remain to strengthen regulatory oversight and improve compliance. Many sponsors still find the reporting interface complex and time-consuming, indicating a need for a more intuitive, streamlined submission platform with automated validation tools to flag incomplete or inconsistent data before submission.^20^ Automation features in the registry could reduce manual validation, while also enhancing safety. These systems could identify study stopping rules and requirements for convening data safety monitoring boards. Moreover, ClinicalTrials.gov could leverage common electronic data capture systems to develop extensions for more robust and timely results submissions.

However, the above solutions require sufficient funding, which remains a hurdle. A 2021 audit inside the NIH revealed that only one compliance officer was monitoring some $4□billion in extramural grants for timely trial registration and reporting, a stark mismatch of staffing to responsibilities.^21^ On the FDA side, oversight of clinical trial results reporting is similarly limited: although the FDA holds the authority to impose civil money penalties (up to around $10,000 per day) for non-compliance, evidence shows no recorded fines have been levied, and the agency lacks a comprehensive internal tracking system for all relevant trials or inspections ^22^. An Office of Inspector General (OIG) report found that the FDA could only inspect about 1□% of trial sites over a multi-year period due to limited resources.^23^ Collectively, these data signal a structural mismatch: rapidly increasing volumes of trial data and user obligations are outpacing the staffing, technological infrastructure, and enforcement capacity at NIH and FDA, contributing to persistent delays and low compliance rates.

## Conclusion

Despite a high rate of clinical trial results submissions, we found widespread delays in the availability of results from antimicrobial clinical trials. Although ACTs outperform pACTs, both groups fall short of federal reporting requirements. Given the urgency of antimicrobial resistance, delayed reporting restricts the evidence base needed for safe and effective treatment. Improving monitoring and simplifying reporting processes could meaningfully enhance compliance. Our findings may inform regulatory strategies in other countries facing similar transparency challenges.

## Acknowledgments

We thank Brix Kowalski for helpful discussions and for his contributions to the conceptualization of this work.

## Data availability

The protocol, data and code can be found here: https://osf.io/zp8ug

## Protocol Deviations

The preregistered protocol differed from the final study in several ways. First, we narrowed the publication search to only include trials with missing results on ClinicalTrials.gov, given the limited resources to manually search all 2,525 NCTs. Second, we updated the scope of the analysis to remove the Interrupted Time Series (ITS) and prospective registration. Although we initially planned to include an ITS, something we had discussed extensively and were enthusiastic about, subsequent evaluation showed that the approach was not well suited to our dataset. The available sample was relatively small and imbalanced over time, several competing model specifications produced unstable estimates, and most importantly, limiting the analysis to antimicrobial agents would have introduced substantial bias in assessing whether FDAAA improved trial reporting. Third, we removed the “Similar-to-ACTs” category to align the terminology with existing research. We referred to the *ClinicalTrials*.*gov Protocol Registration and Results System User’s Guide* to revise the pACT designation as trials with a primary completion date on or after January 2008 and a study start date before January 18, 2017. The study cohort included trials with primary completion dates between May 1, 2013 and May 1, 2023, meaning the pACTs in our analysis had primary completion dates after May 1, 2013 and study start dates before January 18, 2017. Fourth, we modified the data verification by having two individuals (MC) and (AW) perform the data analysis. Another individual (MS) examined the results with a code. The preregistered protocol specified that inter-rater reliability would be assessed using Cohen’s kappa on a random 10% sample of extracted data, with a predefined threshold of 0.8 for acceptable agreement. This assessment was not conducted as planned. This and other minor deviations were due to resource and feasibility constraints related to the scope of manual data extraction.

## References

1. Murray CJL, Ikuta KS, Sharara F, et al. Global burden of bacterial antimicrobial resistance in 2019: a systematic analysis. The Lancet. 2022;399(10325):629–655. doi:10.1016/S0140-6736(21)02724-0

2. Jernigan JA, Hatfield KM, Wolford H, et al. Multidrug-Resistant Bacterial Infections in U.S. Hospitalized Patients, 2012–2017. N Engl J Med. 2020;382(14):1309–1319. doi:10.1056/NEJMoa1914433

3. Antimicrobial resistance. Accessed November 25, 2025. https://www.who.int/news-room/fact-sheets/detail/antimicrobial-resistance

4. Van Noorden R. Report disputes benefit of stockpiling Tamiflu. Nature. Published online April 10, 2014. doi:10.1038/nature.2014.15022

5. Gupta YK, Meenu M, Mohan P. The Tamiflu fiasco and lessons learnt. Indian J Pharmacol. 2015;47(1):11–16. doi:10.4103/0253-7613.150308

6. PRS User’s Guide | ClinicalTrials.gov. Accessed November 26, 2025. https://clinicaltrials.gov/submit-studies/prs-help/user-guide

7. 4.1.3 Clinical Trials Registration and Reporting in ClinicalTrials.gov Requirement. Accessed November 25, 2025. https://grants.nih.gov/grants/policy/nihgps/html5/section_4/4.1.3_clinical_trials_registration_and_reporting_in_clinicaltrials.gov_requirement.htm

8. EBM DataLab. FDAAA TrialsTracker. February 16, 2018. Accessed November 25, 2025. https://fdaaa.trialstracker.net/

9. DeVito NJ, Bacon S, Goldacre B. Compliance with legal requirement to report clinical trial results on ClinicalTrials.gov: a cohort study. The Lancet. 2020;395(10221):361–369. doi:10.1016/S0140-6736(19)33220-9

10. Pellat A, Boutron I, Ravaud P. Availability of results of trials studying pancreatic adenocarcinoma over the past 10 years. Oncologist. Published online August 2022:oyac156.

11. Cho J, Xu Q, Wong CH, Lo AW. Predicting clinical trial duration via statistical and machine learning models. Contemp Clin Trials Commun. 2025;45:101473. doi:10.1016/j.conctc.2025.101473

12. Kao J, Ross JS, Miller JE. Transparency of Results Reporting in Cancer Clinical Trials. JAMA Netw Open. 2023;6(8):e2328117. doi:10.1001/jamanetworkopen.2023.28117

13. Mughal Z, Luechtefeld T, Cashman JC, Tidmarsh GF. Sponsor-Level Compliance with ClinicalTrials.gov Reporting Requirements: A Comprehensive Analysis. J Acad Public Health. Published online September 10, 2025. Accessed November 25, 2025. https://publichealth.realclearjournals.org/research-articles/2025/09/sponsor-level-compliance-with-clinicaltrials-gov-reporting-requirements-a-comprehensive-analysis/

14. El Emam K, Jonker E, Sampson M, Krleža-Jerić K, Neisa A. The Use of Electronic Data Capture Tools in Clinical Trials: Web-Survey of 259 Canadian Trials. J Med Internet Res. 2009;11(1):e8. doi:10.2196/jmir.1120

15. Mughal Z, Fu R, Luechtefeld T, et al. Compliance with Results Reporting at ClinicalTrials.gov Before and After the 2017 FDAAA Final Rule: A Comparative Analysis. J Acad Public Health. Published online January 30, 2025. Accessed November 25, 2025. https://publichealth.realclearjournals.org/research-articles/2025/01/compliance-with-results-reporting-at-clinicaltrials-gov-before-and-after-the-2017-fdaaa-final-rule-a-comparative-analysis/

16. Nilsonne G, Wieschowski S, DeVito NJ, et al. Results reporting for clinical trials led by medical universities and university hospitals in the nordic countries was often missing or delayed. J Clin Epidemiol. 2025;181. doi:10.1016/j.jclinepi.2025.111710

17. Commission Decision (EU) 2021/1240 of 13 July 2021 on the Compliance of the EU Portal and the EU Database for Clinical Trials of Medicinal Products for Human Use with the Requirements Referred to in Article 82(2) of Regulation (EU) No 536/2014 of the European Parliament and of the Council (Text with EEA Relevance). Vol 275.; 2021. Accessed November 25, 2025. http://data.europa.eu/eli/dec/2021/1240/oj

18. Clinical trials - Regulation EU No 536/2014 - Public Health. November 25, 2025. Accessed November 25, 2025. https://health.ec.europa.eu/medicinal-products/clinical-trials/clinical-trials-regulation-eu-no-5362014_en

19. Bakhit M, Jones M, Baker J, et al. Reporting of adverse events, conflict of interest and funding in randomised controlled trials of antibiotics: a secondary analysis. BMJ Open. 2021;11(7):e045406. doi:10.1136/bmjopen-2020-045406

20. Tse T, Fain KM, Zarin DA. How to avoid common problems when using ClinicalTrials.gov in research: 10 issues to consider. BMJ. 2018;361:k1452. doi:10.1136/bmj.k1452

21. Bruckner T. NIH blocks projects of two researchers who fail to make clinical trial results public. TranspariMED. May 11, 2023. Accessed November 25, 2025. https://www.transparimed.org/single-post/gao-audit-nih

22. Commissioner O of the. Civil Money Penalties Relating to the ClinicalTrials.gov Data Bank. November 5, 2024. Accessed November 25, 2025. https://www.fda.gov/regulatory-information/search-fda-guidance-documents/civil-money-penalties-relating-clinicaltrialsgov-data-bank

23. OIG Releases Report of FDA’s Oversight of Clinical Trials, Concludes Improvement of Information Systems and Processes is Needed. Office of Inspector General | Government Oversight | U.S. Department of Health and Human Services. September 28, 2007. Accessed November 25, 2025. https://oig.hhs.gov/newsroom/news-releases-articles/oig-releases-report-of-fdas-oversight-of-clinical-trials-concludes-improvement-of-information-systems-and-processes-is-needed/

